# Dry eye disease: A Canadian quality of life and productivity loss survey

**DOI:** 10.1101/2020.10.07.20207225

**Authors:** Clara C. Chan, Setareh Ziai, Varun Myageri, James G. Burns, C. Lisa Prokopich

## Abstract

**Aim:** To capture the direct and indirect cost estimates of dry eye disease (DED), stratified by disease severity, in patients from Canada and to understand the impact of DED on quality of life (QoL) in this group.

**Methods:** A prospective, multi-centre, observational, cross-sectional study was conducted at six optometry and ophthalmology sites across Canada. Eligible patients completed a 20-minute survey on demography, general health, disease severity, QoL, and direct and indirect costs.

**Results:** A total of 151 patients participated in the study and 146 were included in the analysis. Mean (standard deviation [SD]) age was 49.8 (11.4) years and most patients were female (89.7%). DED was considered moderate or severe by 19.2% and 69.2% of patients, respectively. Sjögren’s syndrome was reported by 8.2% of patients. Total mean annual costs of DED were $24,331 (Canadian dollars [CAD]) per patient and increased with disease severity. Mean (SD) indirect costs for mild, moderate, and severe disease were $5,961 ($6,275), $16,525 ($11,607), and $25,485 ($22,879), respectively. Mean (SD) direct costs were $958 ($1,216), $1,303 ($1,574), and $2,766 ($7,161), respectively. QoL scores were lowest in patients with Sjögren’s syndrome and those with severe DED.

**Conclusions:** This study provides important insights into the negative impact of DED in a Canadian setting. Patients with severe DED reported higher direct and indirect costs and lower QoL compared with those with mild or moderate disease. Increased costs and poorer QoL were also evident for patients with DED plus Sjögren’s syndrome versus DED alone.

## INTRODUCTION

Dry eye disease (DED) is a complex, multifactorial disorder of the periocular tear film that may result in damage to the ocular surface and is associated with symptoms of ocular discomfort.^1^ Globally, the estimated prevalence of DED ranges between 5%–50%.^2^ In Canada, the prevalence of DED has been estimated at 21%.^3^

Diagnosis of DED should consider symptomatology, visual disturbance, tear film stability and composition, tear volume, damage and/or inflammation of the ocular surface, and eyelid aspects (e.g., blepharitis, lid wiper epitheliopathy).^4,5^ In particular, the major cause of DED needs to be recognized before treatment plans can be developed.

The Tear Film and Ocular Surface Society (TFOS) Dry Eye Workshop (DEWS) II proposed a step-wise treatment approach with follow-up to monitor signs and symptoms of DED, in order to ensure that other comorbid ocular surface diseases are also treated appropriately.^1,6^ Initially, DED is often treated with ocular lubricants. If this approach proves inadequate, prescribed topical drugs (e.g., antibiotics, corticosteroids, secretagogues, and immunomodulatory or lymphocyte function-associated antigen-1 antagonist drugs) or oral antibiotics can be considered. Additional options include overnight chamber devices and tear conservation measures.^6^ Where treatment remains ineffective, the next step is to use serum eye drops, therapeutic contact lenses, or oral secretagogues. If the above measures fail to control the disease, further options include lacrimal duct occlusion or longer-term topical corticosteroids (monitored closely to minimize potential adverse side effects such as cataract formation and glaucoma).^6^

DED can incur very high socioeconomic costs, both direct (e.g., medical fees, prescribed drugs) and indirect (e.g., unemployment, work absenteeism, presenteeism [at work but impaired productivity], over-the-counter drugs).^7^ Indirect costs comprise most of the overall economic burden. For example, in the United States (US), the mean average indirect cost to society was US $55.4 billion compared with US $3.8 billion for direct costs.^8^ Furthermore, DED can place a substantial burden on the quality of life (QoL) of patients, impacting their physical, social, and psychological wellbeing, and affecting their workplace productivity.^7,9^ For example, blurred and/or fluctuating vision may restrict activities of daily living such as reading, driving, watching television, and using smartphones.^7^

The physical impact of DED pain can be likened to a type of chronic pain syndrome, negatively affecting patients’ psychological and physical well being and QoL.^7^ Several studies, as reviewed by McDonald et al.,^9^ have demonstrated the impact that DED has upon QoL. However, there are few publications describing the impact of DED upon socioeconomic costs and QoL in Canada.^2,3,10^ The present study aimed to contribute to the literature on DED by capturing the direct and indirect cost estimates of DED, stratified by the severity of DED, and to understand the impact upon QoL for Canadian patients with DED.

## MATERIAL AND METHODS

### Study design

This was a prospective, multi-centre, observational, cross-sectional study conducted at six optometry and ophthalmology sites across Canada. The target population was patients with moderate to severe DED and the study was designed to include 10%, 45%, and 45% of patients with mild, moderate, or severe DED, respectively. The study was performed in accordance with the Health Insurance Portability and Accountability Act 1996 (HIPAA) regulations and the principles of the Declaration of Helsinki. The study was approved by Advarra central institutional review board (IRB). Local IRB and independent ethics committee approvals were obtained according to individual site requirements. Written informed consent was obtained from all patients.

### Patients

Investigators recruited patients for the study according to their diagnosis of DED. Eligible patients were aged 18 to 64 years and were required to meet all of the following criteria: a current diagnosis of mild-to-severe DED made by an accredited healthcare professional; a routine visit to a recruiting healthcare professional; symptoms of dry eye for at least one year before the date of recruitment; literacy in English, ability to read and complete surveys in English, and ability to read, understand, and sign the informed consent form on a voluntary basis. Patients were excluded if they were already participating in another clinical trial.

### Objectives

The two primary objectives of the study were to describe the direct out-of-pocket costs and indirect costs (absenteeism and presenteeism) attributable to DED, and to determine the QoL impact attributable to DED. The four secondary objectives comprised the above analyses conducted in groups stratified by DED severity (mild, moderate, or severe) and by Sjögren’s syndrome status (presence or absence of disease).

### Data collection and assessments

All patients who provided consent were asked to complete a survey lasting approximately 20 minutes (on their own or with help) during their routine optometrist/ophthalmologist visit. Study data were collected from the survey, which was composed of six sections: demographic data; general health data; DED severity data; QoL data; indirect costs; and direct costs. Demographic data (age, sex, ethnicity, education, employment status) and clinical characteristics data (smoking status, contact lens use, screen exposure, DED treatment, DED risk factors, DED duration, ocular and non-ocular comorbidities) over the past 12 months were collected. Patient-reported DED severity and presence or absence of Sjögren’s syndrome were captured. Current DED severity was assessed according to how patients rated their discomfort with eye dryness using the Eye Dryness Score Visual Analog Scale (EDS VAS; score range 0–100 [no discomfort to maximum discomfort]). Scores of <40 were considered mild, 40 to <60 moderate, and ≥60 severe DED. DED severity was also classed as mild, moderate, or severe at the discretion of the patient’s physician. Current QoL was evaluated according to the National Eye Institute Visual Function Questionnaire (NEI VFQ 25; score range 0–100, with lower scores indicating lower QoL).^11^ Overall QoL scores were calculated by averaging vision-targeted subscale scores, excluding general health.

All costs are reported in 2018 Canadian Dollars (CAD). Annual indirect costs were calculated based upon each patient’s annual salary and their Work Productivity and Activity Impairment (WPAI) questionnaire^12^ scores. The WPAI questionnaire comprises questions on how many hours at work were missed due to DED and to what extent DED limited work productivity and non-work activities. Annual indirect costs were inferred using a patient’s highest level of education acquired, and their age and sex, based upon the 2016 Canadian census from Statistics Canada^13^ and adjusted to 2018 levels using the Bank of Canada consumer price index inflation calculator.^14^ Annual direct costs were calculated using patient-reported resource utilization and out-of-pocket expenses for the past 3–24 months. Five categories of treatments were used to assess out-of-pocket costs to DED patients: ocular lubricants, cyclosporine, punctal plugs, optometrist or ophthalmologist visits, and nutritional supplements. A patient’s direct annual cost was the sum of their annualized cost types. Population and subgroup direct costs were calculated using the average of each patient’s direct annual cost values.

### Sample size and statistical analysis

The study aimed to recruit approximately 150 patients from six sites over six months; assuming a standard deviation (SD)-to-mean ratio of 0.14, this would give a relative precision in the primary analysis of up to 7%, with the smallest cohort estimated to contain 15 surveys. All data were analyzed descriptively. Summary statistics for categorical variables included frequency and percentage of each category or modality. Summary statistics for continuous variables are reported as mean and SD. Reported outcomes are composed of at least five patients to preserve patient confidentiality.

## RESULTS

### Patients

In total, 151 patients from six Canadian optometrist and/or ophthalmologist sites consented to participate and the study lasted for seven months (from August 2018 to March 2019). Data from 146 patients were analyzed (**Figure 1**). Data from the remaining five patients were excluded from the analysis, mainly because they had not experienced DED symptoms for at least one year prior to recruitment. Demographic characteristics are shown in **Table 1**. The mean (SD) age was 49.8 (11.4) years. Most patients were female (89.7%), of whom almost half (62/131 [47.3%]) were aged ≥55 years. The majority of patients were Caucasian (71.2%), followed by East Asians (17.8%), and there were only a few patients with mixed ethnicities (2.1%). Most patients (71.9%) held a college degree or a higher qualification, with approximately half of the group (45.9%) educated to university degree level (at or above bachelor level). The remaining patients (28.1%) did not hold a college or higher degree. Most patients were employed (65.1%) and only 2.1% were on disability leave as a result of DED. The mean (SD) annual income was $68,781 ($22,983), with almost half the patients (49.3%) earning >$40,000 but ≤$60,000.

**Figure 1.**
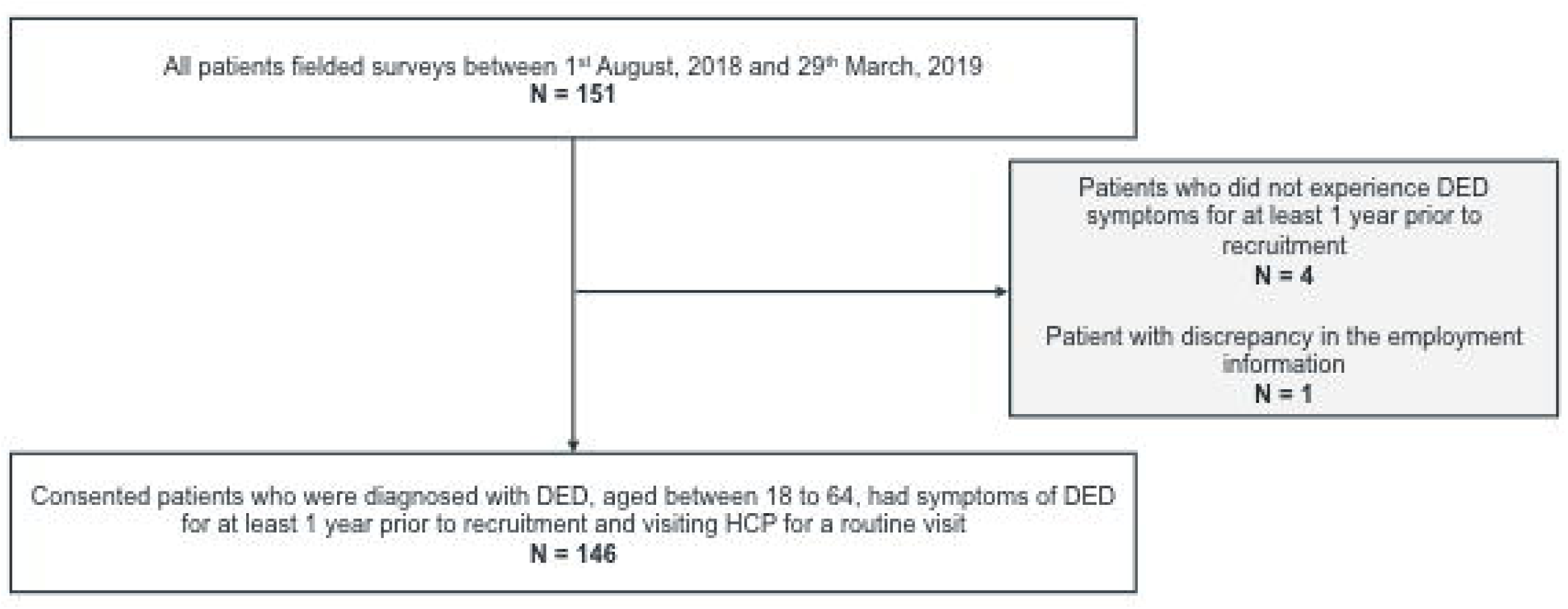

**Table 1.** Patient demographics.

| All values are n (%) unless stated otherwise | Patients<br>N=146 |
| --- | --- |
| Mean (SD) age, years | 49.8 (11.4) |
| Female | 131 (89.7) |
| Age group, years |  |
| 15–24 | 1 (0.7) |
| 25–34 | 19 (13.0) |
| 35–44 | 29 (19.9) |
| 45–54 | 35 (24.0) |
| 55–64 | 62 (42.5) |
| Ethnicity<sup>a</sup> |  |
| Caucasian | 104 (71.2) |
| East Asian | 26 (17.8) |
| South Asian | 8 (5.5) |
| Latino/Hispanic | 3 (2.1) |
| African | 3 (2.1) |
| Middle Eastern | 2 (1.4) |
| Caribbean | 2 (1.4) |
| Other<sup>b</sup> | 4 (2.7) |
| Highest level of education |  |
| University certificate, diploma, or degree |  |
| Above bachelor level | 22 (15.1) |
| At bachelor level | 45 (30.8) |
| Below bachelor level | 4 (2.7) |
| College, CEGEP, or other non-university certificate or diploma | 34 (23.3) |
| Trades |  |
| Certificate of apprenticeship or qualification | 2 (1.4) |
| Certificate or diploma (excluding certificate of apprenticeship) | 9 (6.2) |
| High school diploma or equivalency certificate | 26 (17.8) |
| No certificate, diploma, or degree | 4 (2.7) |
| Employment status |  |
| Employed | 95 (65.1) |
| Disability leave |  |
| Due to DED | 3 (2.1) |
| Other reason | 9 (6.2) |
| I do something else | 6 (4.1) |
| Unemployed |  |
| Job seeking | 9 (6.2) |
| Not job seeking | 9 (6.2) |
| Retired / pre-pension plan | 15 (10.3) |
| Income, CAD |  |
| Mean (SD) | \$68,781 (\$22,983) |
| Median (Q1, Q3) | \$58,755 (\$50,513, \$87,415) |
| Annual income, CAD |  |
| ≤\$40,000 | 4 (2.7) |
| >\$40,000–≤\$60,000 | 72 (49.3) |
| >\$60,000–≤\$80,000 | 24 (16.4) |
| >\$80,000–≤\$100,000 | 32 (21.9) |
| >\$100,000 | 14 (9.6) |

<sup>a</sup>As 3 patients were of mixed ethnicity the total does not add up to 100%.
<sup>b</sup>Includes 1 Filipino; the remaining 3 patients did not specify their ethnicity.
CAD, Canadian dollars; CEGEP, College of General and Vocation Education; DED, dry eye disease; SD, standard deviation.

Clinical characteristics are shown in **Table 2**. Most patients (64.4%) had never smoked, 29.5% had smoked previously, and only 6.2% were current smokers. A minority of patients used contact lenses (13.7%). Most patients (82.3%) spent more than three hours per day looking at screens, with 24.7% looking at screens for at least seven hours per day. DED was considered to be moderate or severe by 19.2% and 69.2% of patients, respectively; however, investigators considered DED to be moderate or severe in 46.7% and 41.1% of patients, respectively. The analysis population only included patients who had been diagnosed with DED for at least one year: 90 patients (61.6%) had experienced DED for 1–5 years and the remainder had a disease duration of ≥6 years.

**Table 2.** Clinical characteristics.

| **All values are n (%) unless stated otherwise** | **Patients**<br>**N=146** |
| --- | --- |
| Smoking status |  |
| Previous smoker | 43 (29.5) |
| Current smoker | 9 (6.2) |
| Never smoker | 94 (64.4) |
| Pregnant |  |
| No | 145 (99.3) |
| Unknown | 1 (0.7) |
| Use contact lenses | 20 (13.7) |
| Screen exposure, hours |  |
| 0-<1 | 10 (6.8) |
| 1-2 | 16 (11.0) |
| 3-4 | 48 (32.9) |
| 5-6 | 36 (24.7) |
| 7-15 | 36 (24.7) |
| Current type of treatment |  |
| Preserved | 33 (22.6) |
| Non-preserved | 52 (35.6) |
| Prescription eye drops | 73 (50.0) |
| Specialty drugs | 11 (7.5) |
| Compounded ointments | 12 (8.2) |
| Gel | 21 (21.2) |
| Other | 39 (26.7) |
| None | 8 (5.5) |
| Concomitant medication |  |
| Multivitamins | 59 (40.4) |
| Antidepressants | 22 (15.1) |
| Oral contraception | 11 (7.5) |
| Anti-anxiety | 8 (5.5) |
| HRT | 6 (4.1) |
| Antihistamines | 5 (3.4) |
| Botox<sup>®</sup> | 4 (2.7) |
| Anticholinergics | 2 (1.4) |
| Beta blockers | 2 (1.4) |
| Diuretics | 2 (1.4) |
| Other<sup>a</sup> | 66 (45.2) |
| None | 32 (21.9) |
| Unknown | 1 (0.7) |

| All values are n (%) unless stated otherwise | Patients<br>N=146 |
| --- | --- |
| Sjögren's syndrome |  |
| No | 122 (83.6) |
| Yes | 12 (8.2) |
| Don't know | 11 (7.5) |
| Unknown | 1 (0.7) |
| Diagnosis risk factors |  |
| Thyroid disorder | 22 (15.1) |
| Autoimmune diseases<sup>b</sup> | 20 (13.7) |
| Chronic pain | 20 (13.7) |
| Migraines | 20 (13.7) |
| Mental health conditions<sup>c</sup> | 19 (13.1) |
| Irritable bowel syndrome | 17 (11.6) |
| Rosacea | 12 (8.2) |
| Diabetes | 8 (5.5) |
| Fibromyalgia | 7 (4.8) |
| HSCT or GvHD | 2 (1.4) |
| Androgen deficiency | 1 (0.7) |
| Gout | 1 (0.7) |
| Vitamin A deficiency | 1 (0.7) |
| Other | 31 (21.3) |
| None | 47 (32.2) |
| Unknown | 2 (1.4) |
| Patient-reported severity of DED |  |
| Mild | 17 (11.6) |
| Moderate | 28 (19.2) |
| Severe | 101 (69.2) |
| DED duration, years |  |
| 1–5 | 90 (61.6) |
| 6–10 | 28 (26.0) |
| 10–35 | 18 (12.3) |
| Ocular comorbidities |  |
| Seasonal allergies with itchy eyes | 32 (21.9) |
| Meibomian gland dysfunction | 27 (18.5) |
| Prior conjunctivitis (pink eye) | 13 (8.9) |
| Glaucoma | 2 (1.4) |
| Surfer's eye (pterygium) | 2 (1.4) |
| Keratoconus +/- corneal collagen crosslinking | 1 (0.7) |
| Other | 11 (7.5) |
| None | 77 (52.7) |
| Ocular surgery |  |
| Refractive eye surgery<sup>d</sup> | 23 (15.8) |
| Eyelid surgery | 8 (5.5) |
| Cataract surgery | 6 (4.1) |
| Glaucoma surgery | 1 (0.7) |
| Other | 14 (9.6) |
| None | 99 (67.8) |
| Unknown | 1 (0.7) |

<sup>a</sup>Including hypertension, antidiabetic, and analgesic medications.
<sup>b</sup>For example, Sjögren's syndrome, rheumatoid arthritis, lupus.
<sup>c</sup>For example, mood, depression, anxiety.
<sup>d</sup>Including laser-assisted in situ keratomileusis or photorefractive keratectomy
DED, dry eye disease; GvHD, graft vs. host disease; HRT, hormone replacement therapy; HSCT, hematopoietic stem cell transplantation.

An absence of ocular comorbidities and/or surgeries/injections accounted for 52.7% and 67.8% of patients, respectively. The remainder reported a variety of ocular comorbidities, of which the most frequent were seasonal allergies with itchy eyes (21.9%) and Meibomian gland dysfunction (18.5%). The most frequent ocular surgery was refractive eye surgery, which had been conducted in 15.8% of patients. The most frequently used treatment reported by patients was eye drops (preserved, 22.6%; non-preserved, 35.6%). Concomitant medications included multi-vitamins, which were being taken by 40.4% of patients, and anti-depressants (used by 15.1% of patients). A total of 21.9% of patients were not taking any concomitant medications.

Twelve patients (8.2%) reported having Sjögren’s syndrome, 122 (83.6%) did not have the condition, and the status was unknown for 12 patients (8.2%). Among other diagnostic risk factors, thyroid disorders were reported by 15.1% of patients whereas 32.2% did not report any diagnostic risk factors.

### Annual costs of DED

The total annual costs of DED per patient were on average $24,331 (**Figure 2**); the majority were related to the indirect costs of DED presenteeism (79.3%). The mean (SD) patient-reported indirect cost attributable to DED was $21,052 ($20,812)/year (**Figure 2**). Mean (SD) indirect costs of presenteeism and absenteeism were $19,304 ($18,916)/year and $2,702 ($11,028)/year, respectively. The mean (SD) total patient-reported direct cost attributable to DED was $2,324 ($6,159)/year (**Figure 2**).

**Figure 2.**
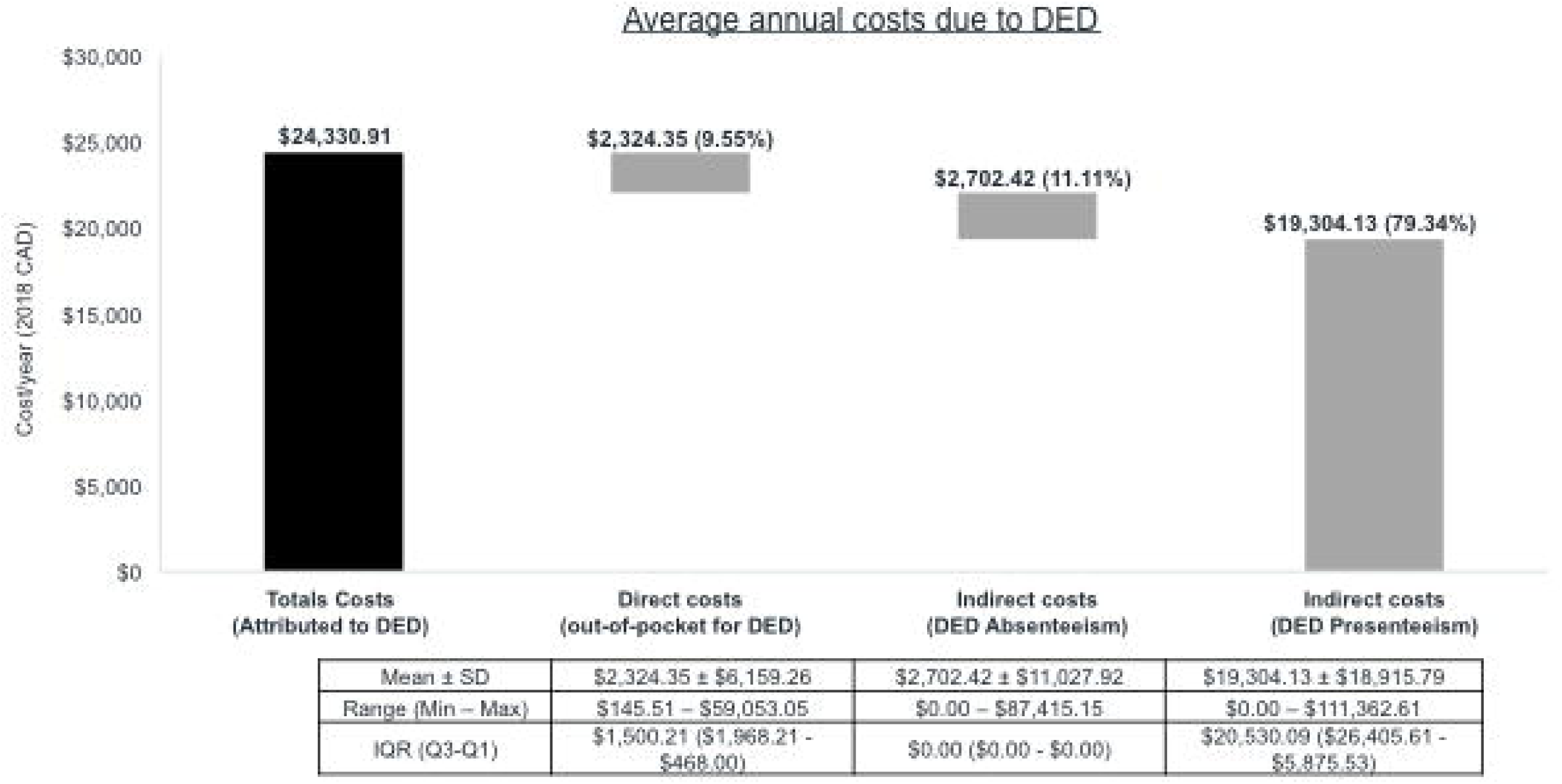

When average annual costs were stratified by DED severity, the mean (SD) indirect costs of absenteeism and presenteeism attributable to severe DED totaled $25,485 ($22,879)/year, representing an increase of 54% versus the costs of moderate DED ($16,525 [$11,607]/year) and 328% versus mild DED ($5,961 [$6,275]/year). Direct out-of-pocket costs per year were also highest for severe DED (mean [SD]: $2,766 [$7,161]), representing an increase of 112% versus the costs of moderate DED ($1,303 [$1,574]) and 189% versus mild DED ($958 [$1,216]).

The mean (SD) indirect costs per year of absenteeism and presenteeism attributable to Sjögren’s syndrome totalled $41,094 ($15,720), representing an increase of 132% versus the costs for patients without Sjögren’s syndrome ($17,694 [$17,153]). Direct out-of-pocket costs were also higher for Sjögren’s syndrome (mean [SD]: $2,689 [$2,430]), representing an increase of 22% versus the costs for patients without Sjögren’s syndrome ($2,203 [$6,660]).

### Impact of DED on QoL

The total mean (SD) VFQ-25 QoL score was 77 (16) (**Figure 3**). Ocular pain had the lowest score of all subscales (mean [SD] score 52 [22]), indicating that it had the greatest negative effect on QoL. DED also impacted mental health (66 [26]), role difficulties (at work and for other activities, 68 [26]), and driving (75 [19]). On average, patients with severe DED reported a lower total VFQ-25 QoL score (mean 72 [SD 16]) than patients with moderate (87 [7]) or mild (90 [7]) DED. The mean (SD) total VFQ-25 QoL score was lower in patients with Sjögren’s syndrome compared with those without the condition (60 [14] vs. 79 [15]), respectively.

**Figure 3.**
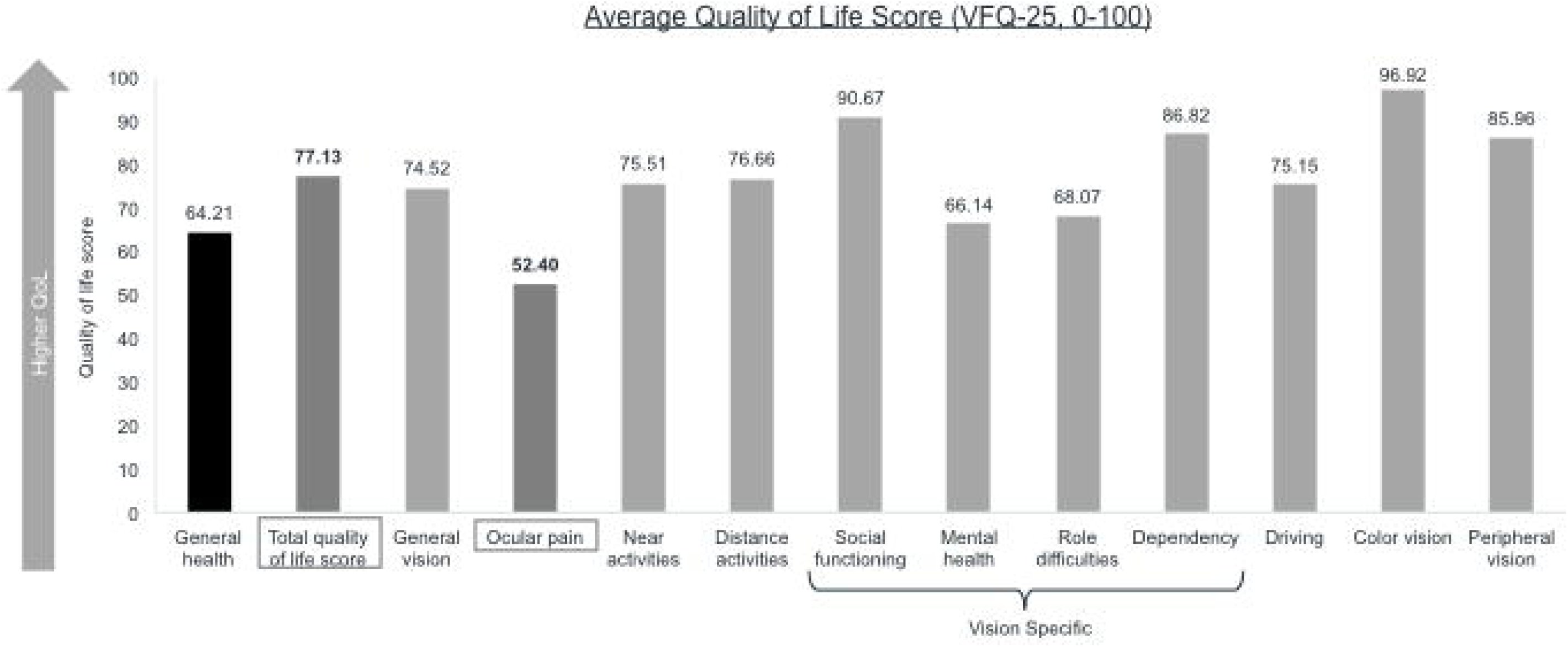

## DISCUSSION

This study provides an analysis of the economic burden and effects on QoL in Canadian patients with DED. Patients with severe DED reported lower QoL, higher medical fees, and impaired work productivity compared with mild and moderate cases. Annual indirect costs increased from a mean (SD) of $5,961 ($6,275) in mild DED to $25,485 ($22,879) in severe DED. Direct costs followed a similar pattern, with a mean (SD) of $958 ($1,216)/year in mild DED rising to $2,766 ($7,161)/year in severe DED. Patients with Sjögren’s syndrome also incurred higher costs than those without Sjögren’s syndrome. The mean total QoL score was 77, with ocular pain impacting patients’ QoL the most (mean NEI VFQ-25 score 52); DED also negatively affected patients’ mental health, role difficulties, and driving. Unsurprisingly, QoL was worse in patients with severe DED versus those with mild/moderate disease, and in those with versus those without Sjögren’s syndrome.

Much of the previous literature has evaluated the costs and burden of DED from a payers’ or healthcare systems perspective, and has found that DED confers a substantial economic burden. In a systematic literature review, the majority of economic data was found to be from the US (9/12 articles), and health-related QoL data were predominantly from Europe (11/20 articles) and the US (8/20 articles).^9^ McDonald et al. found that indirect costs comprised the largest proportion of overall DED costs, due to significant loss in work productivity.^9^ In the US, for example, indirect costs were US $11,302/year per patient (with the overall burden to society being US $55 billion), and the numbers of days lost per year for mild, moderate, and severe DED were 91, 95, and 128, respectively.^8^ None of the 12 economic burden articles included Canadian data, although one article reported Canadian QoL data.

Our research provides additional insights into the burden of DED in a Canadian setting, taking the patients’ direct costs into account and estimating indirect costs to society. We have shown that in Canada, indirect costs comprise the largest proportion of overall costs compared with direct costs. We found that DED impacts both indirect and direct costs. In addition, direct costs increased with disease severity. Our data are supported by an analysis of 2,171 US patients, where indirect versus direct costs were estimated as US $11,302/year versus $783/year per patient.^8^ Likewise, direct costs increased with disease severity (US $678/year for mild DED, US $771 for moderate DED, and US $1,267 for severe DED).^8^

Similar to our data, in which patients ranked ocular pain as the lowest favorable item in terms of QoL, impaired QoL in patients with active primary Sjögren’s syndrome is mostly caused by ocular pain and dryness.^15^ In the US, the mean ocular pain subscale score was significantly lower for patients with moderate-to-severe DED compared with patients with milder DED.^16^ Additionally, assessment of the relative burden of DED in the US showed that DED consistently caused bodily pain (effect size −0.08) and decreased role-physical (defined as limitations due to physical problems, −0.07), and vitality scores (−0.11) when compared to people without DED. However, the difference was only clinically significant for moderate or severe DED.^17^ QoL was impaired to a greater extent in patients with Sjögren’s syndrome versus those with non-Sjögren’s syndrome DED.^17^ Our data are in agreement with these observations.

While our findings provide important evidence on the economic and QoL burden associated with DED in a Canadian population, they must be interpreted in context. Canada has a distinct climate with the potential to worsen the symptoms of DED as a result of dry heat indoors in the winter, a requirement for air conditioning during the summer, and the effect of spring and fall allergy seasons. As this was a survey-based study, it relied on human recall to evaluate productivity loss, QoL impact and out-of-pocket costs. Recall bias was minimized by using validated recall periods for the VFQ-25 and WPAI questionnaires.^18,19^ Selection bias is a typical limitation of cross-sectional surveys as recruitment of patients with DED who had visited an optometrist/ophthalmologist could lead to an underestimation of productivity losses and their costs (due to patients being too sick to attend a routine visit). An overestimation of losses is also possible because patients with mild DED may not have required a routine visit during the study period. The findings of our study are limited by the sample size and subsequent stratification. Missing or invalid data were minimal, as the investigators checked completed surveys prior to patient discharge. As our analysis was descriptive, and no hypothesis was tested, confounding is not expected to impact our findings.

## CONCLUSIONS

A cross-sectional survey assessed the economic burden and QoL associated with DED in a sample of patients living in Canada. The annual cost of DED averaged $24,330, with indirect costs accounting for 90% of the total. Direct and indirect costs, and the negative impact of DED on QoL, were higher in patients with severe DED compared to those with mild or moderate disease. Sjögren’s syndrome was also associated with higher costs and lower QoL than in patients without Sjögren’s syndrome. This study furthers our understanding of the influence of DED on everyday life for patients with this condition.

## Data Availability

Data are proprietary to IQVIA Solutions and are not publicly available.

## Disclosures

Clara C. Chan has been a consultant for Alcon, Allergan, Bausch+Lomb, Johnson & Johnson, Labtician, Santen, and Shire,* and has received research support from Allergan, Bausch+Lomb, and TearLab.

Setareh Ziai has received grant/research support from Shire* and honoraria/consulting fees from Allergan, Labtician, J&J, Santen, and Shire*.

Varun Myageri was a paid consultant to Shire* in connection with this study.

James G. Burns was an employee of Shire* at the time of the study and is a current employee of Novartis.

C. Lisa Prokopich has been a consultant for Alcon Canada, Innova Medical, Shire*, Novartis, and Bausch+Lomb Canada.

*A Takeda company

## Acknowledgements

The authors would like to thank the Vector Eye Centre, Calgary, Alberta (Jamie Bhamra, Lead Physician; Alison Endsin, Study Coordinator), Rosedale Medical Centre, Toronto, Ontario (Clara C. Chan, Lead Physician; Paul Lam, Study Coordinator), Ivey Eye Institute, London, Ontario (Rookaya Mather, Lead Physician; Bobbi Smuck and Julie Duncan, Study Coordinators), Clarity Eye Institute, Scarborough, Ontario (King Chow, Lead Physician; Heather Coulter, Study Coordinator), Valley Family Optometry (Krista Flynn, Lead Physician; Shyanne Jenkins, Study Coordinator), and Elmsdale Vision Centre, Elmsdale, Nova Scotia (Andrew Webber, Lead Physician; Dolly Chippett and Stacey MacKenzie, Study Coordinators), and the patients who participated in this study. The authors also wish to thank Johanna Mancini, Megan Naeini, and Deepi Minhas of IQVIA for their help in the design, management, and analysis of the study, and Christopher Reaume of Takeda.

## REFERENCES

1. Craig JP, Nichols KK, Akpek EK, et al. TFOS DEWS II Definition and Classification Report. Ocul Surf 2017;15:276–83.

2. Stapleton F, Alves M, Bunya VY, et al. TFOS DEWS II Epidemiology Report. Ocul Surf 2017;15:334–65.

3. Caffery B, Srinivasan S, Reaume CJ, et al. Prevalence of dry eye disease in Ontario, Canada: A population-based survey. Ocul Surf 2019;17:526–31.

4. Wolffsohn JS, Arita R, Chalmers R, et al. TFOS DEWS II Diagnostic Methodology report. Ocul Surf 2017;15:539–74.

5. Pflugfelder SC, Solomon A, Stern ME. The diagnosis and management of dry eye: a twenty-five-year review. Cornea. 2000;19:644–9.

6. Jones L, Downie LE, Korb D, et al. TFOS DEWS II Management and Therapy Report. Ocul Surf 2017;15:575–628.

7. Uchino M, Schaumberg DA. Dry Eye Disease: Impact on Quality of Life and Vision. Curr Ophthalmol Rep 2013;1:51–7.

8. Yu J, Asche CV, Fairchild CJ. The economic burden of dry eye disease in the United States: a decision tree analysis. Cornea 2011;30:379–87.

9. McDonald M, Patel DA, Keith MS, Snedecor SJ. Economic and Humanistic Burden of Dry Eye Disease in Europe, North America, and Asia: A Systematic Literature Review. Ocul Surf 2016;14:144–67.

10. Abetz L, Rajagopalan K, Mertzanis P, Begley C, Barnes R, Chalmers R. Development and validation of the impact of dry eye on everyday life (IDEEL) questionnaire, a patient-reported outcomes (PRO) measure for the assessment of the burden of dry eye on patients. Health Qual Life Outcomes 2011;9:111.

11. RAND. The National Eye Institute 25-Item Visual Function Questionnaire (VFQ-25) Manual 2000. https://www.rand.org/content/dam/rand/www/external/health/surveys_tools/vfq/vfq25manual.pdf.

12. Reilly Associates. WPAI:SHP v2.0 (US English) 2010. http://www.reillyassociates.net/WPAI_SHP.html.

13. 2016 Census of Population - Employment Income Statistics (table) [Internet] Statistics Canada. 2017. http://www12.statcan.gc.ca/census-recensement/2016/dp-pd/dt-td/Rp-eng.cfm?LANG=E&APATH=3&DETAIL=0&DIM=0&FL=A&FREE=0&GC=0&GID=0&GK=0&GRP=1&PID=110647&PRID=10&PTYPE=109445&S=0&SHOWALL=0&SUB=0&Temporal=2017&THEME=123&VID=0&VNAMEE=&VNAMEF.

14. Consumer Price Indexes for Canada [Internet]. Bank of Canada. https://www.bankofcanada.ca/rates/related/inflation-calculator/.

15. Cornec D, Devauchelle-Pensec V, Mariette X, et al. Severe Health-Related Quality of Life Impairment in Active Primary Sjogren’s Syndrome and Patient-Reported Outcomes: Data From a Large Therapeutic Trial. Arthritis Care Res (Hoboken) 2017;69:528–35.

16. Nichols KK, Mitchell GL, Zadnik K. Performance and repeatability of the NEI-VFQ-25 in patients with dry eye. Cornea 2002;21:578–83.

17. Mertzanis P, Abetz L, Rajagopalan K, et al. The relative burden of dry eye in patients’ lives: comparisons to a U.S. normative sample. Invest Ophthalmol Vis Sci 2005;46:46–50.

18. Reilly MC, Zbrozek AS, Dukes EM. The validity and reproducibility of a work productivity and activity impairment instrument. Pharmacoeconomics 1993;4:353–65.

19. Mangione CM, Lee PP, Pitts J, Gutierrez P, Berry S, Hays RD. Psychometric properties of the National Eye Institute Visual Function Questionnaire (NEI-VFQ). NEI-VFQ Field Test Investigators. Arch Ophthalmol 1998;116:1496–504.

